# Historical state compulsory schooling laws and pandemic-era mortality: A quasi-experimental study

**DOI:** 10.1101/2024.02.29.24303564

**Authors:** Whitney Wells, Yea-Hung Chen, Marie-Laure Charpignon, Ah-Reum Lee, Ruijia Chen, Andrew C. Stokes, Jacqueline M. Torres, M. Maria Glymour

## Abstract

**Introduction:** Low educational attainment is associated with increased risk of COVID-19 mortality, but it remains unclear whether the link between education and COVID-19 mortality is causal or due to confounding factors, such as childhood socio-economic status or cognitive skills. To address this question, we evaluated whether older adults’ risk of COVID-19 mortality was associated with historical state-level compulsory schooling laws (CSLs) applicable when they were school-aged. We also evaluated whether that impact was unique to COVID-19 mortality or also applied to all-cause mortality, both before and during the pandemic.

**Methods:** We defined mortality outcomes using US death certificate data from Mar 2019-Dec 2021 for people born in the US before 1964 in three time periods: the year prior to the pandemic (Mar 2019-Feb 2020), pandemic year 1 (Mar 2020-Feb 2021), and pandemic year 2 (Mar-Dec 2021). We determined the population at risk using 2019 American Community Survey PUMS data with population weights, representing 78.7 million individuals born in the US before 1964. We linked individuals to the number of mandatory years of education defined by CSLs specific to their state of birth and years when school age. We estimated intention-to-treat effects of CSLs on mortality using logistic regressions controlling for state-of-birth fixed effects, birth year (linear and quadratic), sex, race, ethnicity, and state-level factors including percent urban, Black, and foreign-born (at age 6) and manufacturing jobs per capita and average manufacturing wages (at age 14).

**Results:** We identified a dose-response relationship between CSLs and mortality. In the first year of the pandemic, people mandated to receive 8 vs 9 (reference) years of education had higher odds of COVID-19 mortality (Odds Ratio [OR]: 1.15; 95% Confidence Interval [CI]: 1.10, 1.19), while those mandated to receive 10 vs 9 (reference) years of education had lower odds of COVID-19 mortality (OR: 0.96; 95% CI: 0.94, 0.98). The association of CSLs with COVID-19 mortality was similar in pandemic years 1 and 2; for all-cause mortality in pandemic years 1 and 2; and for all-cause mortality in the year prior to the pandemic. Results were robust to alternative model specifications.

**Conclusions:** These findings support a causal benefit of education for reduced mortality during the COVID-19 pandemic. State investments in children’s education may have reduced pandemic-era mortality decades later. Our research has implications beyond the pandemic context, as our results suggest the observed relationship mirrors a pre-existing relationship between CSLs and all-cause mortality.

## Introduction

The COVID-19 pandemic underscored inequities in US morbidity and mortality and the importance of social determinants of health.^1–3^ Educational attainment was among the strongest predictors of excess mortality in the first year of the pandemic.^4,5^ For instance, from March 2020 to February 2021, Californians without a high-school diploma experienced over five times the per-capita excess mortality of those with a graduate or professional degree.^5^ Nation-wide examination of county-level COVID-19 rates found the proportion of individuals in a county without a high-school degree was more strongly associated with COVID-19 case and fatality rate than poverty, employment, and housing vacancy.^6^

Educational attainment could influence COVID-19 mortality and all-cause mortality through multiple pathways, such as via occupational exposures, health literacy, access to healthcare institutions, and financial resources.^5^ However, the relationship between education and mortality could in part be confounded by early-life social factors such as parental education or socioeconomic status, or cognitive skills established prior to school–most of which are unmeasured or inconsistently reported in survey data and vital records. Thus, more research is needed to understand whether the relationship between education and mortality is causal or at least partially due to individual-level confounding.

Throughout the first half of the 20th century, the US implemented educational policies with the goal of increasing educational attainment. Among the most important of these were compulsory schooling laws (CSLs) that specified the age at which children were required to enter school and allowed to leave school. During this period, CSLs varied by state. Prior literature has examined the effects of CSLs on a range of health outcomes.^7^ For example, CSLs have been found to impact physical and mental health,^8^ old age mental status,^9^ and cardiovascular outcomes and risk factors.^10^

This research leverages historical variation in CSLs to investigate whether mandates of additional years of education led to a decrease in COVID-19 mortality. To understand whether CSLs had a unique impact on COVID-19 mortality, we compared this to how CSLs affected all-cause mortality, both during the pandemic and before the pandemic. We hypothesized that exposure to more years of state-mandated schooling decades ago would lead to lower all-cause and COVID-19 mortality.

Following Fundamental Cause Theory (FCT), we expected that the mortality benefit of policies aimed at increasing education might vary over time and based on whether there are known strategies to prevent a major cause of death. For example, as knowledge of COVID-19 prevention and treatment grew, including with the introduction of COVID-19 vaccines, we expected inequalities related to education may also grow. FCT suggests that individuals with socio-economic resources that stem from greater educational attainment are able to deploy these resources flexibly to protect themselves in the face of health-related threats, such as via working in occupations that allowed for physical distancing or access to knowledge about health-protective resources (e.g. masks).^11,12^

## Methods

### Population data and measures

We used the 2019 American Community Survey (ACS) Public Use Microdata Sample (PUMS) to estimate the population of individuals who were at risk of death during our time periods.^13^ We used ACS population weights to represent the cohort of Americans satisfying our eligibility criteria. We selected 1-year 2019 data, which sampled households and group quarters to capture roughly one percent of the US population for our population denominator values to get the best estimate of the population at risk of COVID-19 mortality in the year prior to the pandemic.^14^

### Outcome data and measures

Our outcomes were mortality with COVID-19 listed as an underlying cause of death and all-cause mortality. We considered all-cause mortality during the pandemic in part to account for undiagnosed COVID-19 deaths. We used individual-level death records from the National Center for Health Statistics for deaths between March 2019 and December 2021, split into three periods: pre-pandemic (Mar 2019-Feb 2020), pandemic year 1 (Mar 2020-Feb 2021), and pandemic year 2 (Mar 2021-Dec 2021). We defined COVID-19 deaths as those with a primary *International Classification of Diseases, Tenth Revision* diagnostic code of U071 as the underlying cause.

### Study sample

Given the available dates of CSL measures, we restricted our analysis to individuals born before 1964. We excluded individuals born outside the US, or in states without CSLs in this time period (Alaska and Hawaii). ACS performs top-coding on age to prevent de-anonymization of individuals in small groups. ACS calculates a top-code age for each state based on the age distribution, where the age of individuals above the top-code is replaced with a prespecified value. We excluded those top-coded on age given we would not be able to match to the correct CSL based on birth year. The state with the oldest top-code for age corresponds to a birth year of 1926, therefore our data does not include anyone born before 1926. To match the age distribution in the population data, we excluded individuals born before 1926 in the mortality data. Our final sample represented a population of 85.1 million adults born before 1964 (Figure 1).

**Figure 1.**
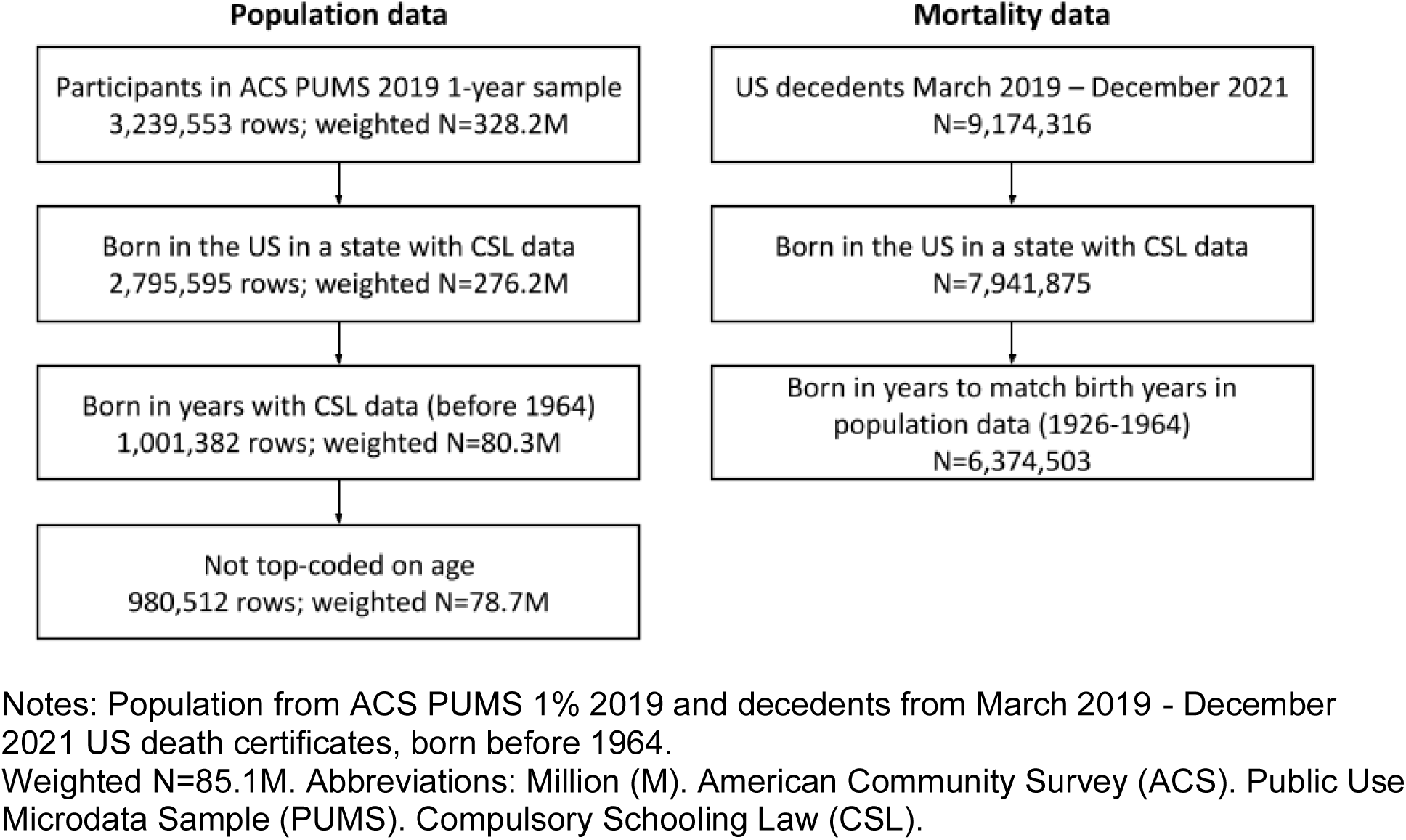
Sample inclusion flow chart.

### Exposure data and measures

Our exposures were state-level mandatory years of education based on CSLs. In our main analysis, we defined the number of mandatory years of education by subtracting mandatory school enrollment age from legal drop-out age. As a supplemental analysis, we defined this number by subtracting mandatory enrollment age from the age at which students were allowed to receive a work permit.^9^ To estimate the schooling measures that would most likely apply to a given individual, we lagged mandatory enrollment age by six years after birth year and lagged drop-out age and work permit age by 14 years. CSL data from 1907-1961 were compiled by Lleras-Muney,^15^ Angrist and Acemoglu,^16^ and Glymour et al.^9,17^ using US federal education reports usually published biennially. Compulsory schooling law measures were filled by carrying forward the most recently recorded measure until a new measure was recorded.^8^

### Covariates

Covariates included both individual-level and state-level characteristics. We adjusted for year of birth (linear and quadratic) to account for secular trends, state of birth fixed effects to account for other state-level factors, and selected time-varying state-level factors, compiled from Statistical Abstracts of the United States and other US Census Bureau reports.^9,18,19^ Time-varying state-level factors included (i) percent urban, percent Black, and percent foreign-born in the respondent’s state of birth when they were 6 years old and (ii) average manufacturing wages and manufacturing jobs per capita in their state of birth when they were 14 years old. Linear interpolation was used to impute characteristics in years between reports. Individual-level sex, race, and ethnicity were included for investigation as potential modifiers of the effect of CSLs on mortality.

## Statistical analysis

### Creation of main analytic dataset

We stacked mortality and population data to create a single dataset representative of individuals within our target population and decedents. Individuals present in the mortality dataset were assigned a death outcome of 1, while individuals present in the population dataset were assigned a death outcome of 0. Notably, some individuals who participated in the 2019 ACS survey might have died during our study period and thus been present in both the mortality dataset (with an outcome of 1) and the population dataset (with an outcome of 0). Our approach does not allow identification of individuals in the ACS who died after 2019; they are thus represented twice in our analytic data set. However, they account for a negligible proportion of the overall susceptible population so the estimates will be quite similar to those that would be generated had they not been double counted. In the stacked dataset, rows from the ACS dataset were assigned the corresponding ACS population weight, while rows from the mortality dataset were assigned a weight of 1. In our analysis of COVID-19 mortality, deaths from any other underlying causes were excluded. Our datasets did not capture changes in state of residence during childhood. Therefore, each individual was linked to the CSLs that corresponded to their year and state of birth, with the assumption that inter-state migration between birth and school-age was minimal. Prior research has demonstrated inter-state migration during this period was relatively low and unrelated to CSL implementation, therefore any measurement error would likely tend to bias results towards the null.^10,15,20^

### Statistical methods

We estimated the effect of CSLs on mortality using multivariate logistic regression. We present odds ratios (OR) of mortality for each level of CSLs, compared to the most common level of mandated school (9 years) as the reference. These estimates can be interpreted as the odds of the specified mortality outcome among people born in a cohort for which their birth state mandated 6, 7, 8, 10, 11, or 12 years of school compared to the odds of mortality among people born in a cohort for which the birth state mandated 9 years of school.

We adjusted for all covariates listed above. In our main analysis, we did not adjust for individual-level factors that may confound the relationship between individual educational attainment and mortality (i.e., sex, race, ethnicity), as these do not confound the effect of CSLs. A conceptual diagram is included as Supplemental Figure 1. In exploratory analyses, we examined potential effect modification by sex, race, and ethnicity (included in the Supplement).

We examined mortality in three time periods. We defined year 1 of the pandemic as Mar 2020 – Feb 2021 and year 2 as Mar 2021 – Dec 2021. At the time of this analysis, final mortality data was not available beyond the end of December 2021. We consider year 1 and year 2 of the pandemic separately given differences in disease dynamics and mortality following the distribution of vaccines. For example, as of March 2021, at least half of adults aged 65+ had received 1 or more doses of a COVID-19 vaccine in most US states.^21^ We also considered the year immediately before the pandemic (Mar 2019 – Feb 2020) to compare our results to pre-pandemic all-cause mortality. In sum, we examined all-cause mortality in all three periods and COVID-19 mortality in pandemic years 1 and 2. To assess the robustness of our findings to this data limitation resulting in a right-truncated year 2 given the last data of final death data, we included a sensitivity analysis specifying alternative time periods in the Supplement.

Analysis was performed in Stata version 17. Code review was performed on all statistical code. PUMS data are publicly available. The use of restricted death certificate data underwent review and approval by NCHS, and data was handled in compliance with the Data Use Agreement. The IRB at the University of California, San Francisco determined that the research did not meet the regulatory definition of human subjects research and did not require formal IRB oversight.

## Results

### Sample characteristics

The weighted sample represented a US population of 78.7 million individuals, with 502,779 COVID-19 deaths and 6.4 million all-cause deaths across the 3 years (Table 1). Compared to non-decedents, decedents were older, and more likely to be male, Black, and have lower educational attainment. Compared to decedents of all-causes, those who died of COVID-19 were similarly aged, but were even more likely to be male, Black, and have lower education. For example, 9.3% of the population had less than a high school diploma as compared to 15.7% of those who died of any cause and 17.6% of those who died of COVID-19.

**Table 1.** Sample characteristics of population and decedents, weighted.

|  | **Susceptible population**<br>(n=78.7M) |  | **Decedents (all causes)**<br>(n=6.4M) |  | **Decedents (COVID-19)**<br>(n=502,779) |  |
| --- | --- | --- | --- | --- | --- | --- |
|  | *N / Mean* | *(% / SD)* | *N / Mean* | *(% / SD)* | *N / Mean* | *(% / SD)* |
| Age | 67.4 | (8.8) | 75.8 | (10.4) | 75.0 | (10.2) |
| Sex |  |  |  |  |  |  |
| Male | 36,707,418 | (46.6%) | 3,311,290 | (51.9%) | 275,419 | (54.8%) |
| Female | 41,997,790 | (53.4%) | 3,063,213 | (48.1%) | 227,360 | (45.2%) |
| Race/Ethnicity |  |  |  |  |  |  |
| White | 65,112,435 | (82.7%) | 5,308,793 | (83.3%) | 386,430 | (76.9%) |
| Black | 8,613,861 | (10.9%) | 795,042 | (12.5%) | 78,474 | (15.6%) |
| Asian | 296,698 | (0.4%) | 11,063 | (0.2%) | 919 | (0.2%) |
| Hispanic | 3,421,597 | (4.3%) | 202,270 | (3.2%) | 29,328 | (5.8%) |
| Mixed/Other | 1,260,617 | (1.6%) | 57,335 | (0.9%) | 7,628 | (1.5%) |
| Reported education |  |  |  |  |  |  |
| <=8th grade | 2,209,088 | (2.8%) | 389,058 | (6.1%) | 37,010 | (7.4%) |
| 9-12th grade, no diploma | 5,082,624 | (6.5%) | 613,098 | (9.6%) | 51,189 | (10.2%) |
| HS diploma/GED | 24,715,801 | (31.4%) | 2,853,916 | (44.8%) | 232,233 | (46.2%) |
| >=Some college | 46,697,695 | (59.3%) | 2,451,884 | (38.5%) | 176,163 | (35.0%) |
| Missing | 0 | (0.0%) | 66,547 | (1.0%) | 6,184 | (1.2%) |
| CSLs based on dropout age | 9.2 | (0.9) | 9.1 | (0.9) | 9.2 | (0.9) |
| CSLs based on work permit age | 8.1 | (1.1) | 7.9 | (1.2) | 7.9 | (1.1) |
| State of birth characteristics, age 6 |  |  |  |  |  |  |
| Percent urban | 66.8 | (16.4) | 60.7 | (18.6) | 60.7 | (18.5) |
| Percent Black | 10.8 | (10.1) | 11.3 | (11.4) | 12.0 | (11.6) |
| Percent foreign | 5.8 | (4.8) | 6.6 | (6.0) | 6.2 | (5.9) |
| State of birth characteristics, age 14 |  |  |  |  |  |  |
| Manufacturing jobs per capita | 68.8 | (25.6) | 70.5 | (30.2) | 69.7 | (30.1) |
| Avg. manufacturing wages | 40765.3 | (8387.2) | 34615.5 | (9046.7) | 34918.7 | (8908.2) |

Notes: Population from ACS PUMS 1% 2019 and decedents from March 2019 - December 2021 US death certificates, born before 1964.
Weighted N=85.1M. Abbreviations: Million (M). American Community Survey (ACS). Public Use Microdata Sample (PUMS). Compulsory Schooling Laws (CSLs). High school (HS). General Education Development (GED).

### Effect of CSLs on COVID-19 mortality

We observed a dose-response relationship between CSLs and COVID-19 mortality. Each additional year of mandatory schooling was associated with lower odds of COVID-19 mortality (Figure 2). In the first year of the pandemic, individuals mandated to complete 8 rather than 9 years of education (reference) had higher odds of COVID-19 mortality (Odds Ratio [OR]: 1.15; 95% Confidence Interval [CI]: 1.10, 1.19). Conversely, individuals mandated to complete 10 rather than 9 years of education had lower odds of COVID-19 mortality (OR: 0.96; 95% CI: 0.94, 0.98). Being mandated to complete 8 (vs 9) years of schooling was also associated with higher odds of COVID-19 mortality during pandemic year 2 (OR: 1.13; 95% CI: 1.09, 1.18), while 10 (vs 9) years was associated with lower odds (OR: 0.97; 95% CI: 0.95, 0.99). Individuals with less than 8 or more than 10 years of mandatory education represent much smaller groups, so estimates are imprecise but the gradient of lower mortality with additional years of mandated schooling was present for both periods.

**Figure 2.**
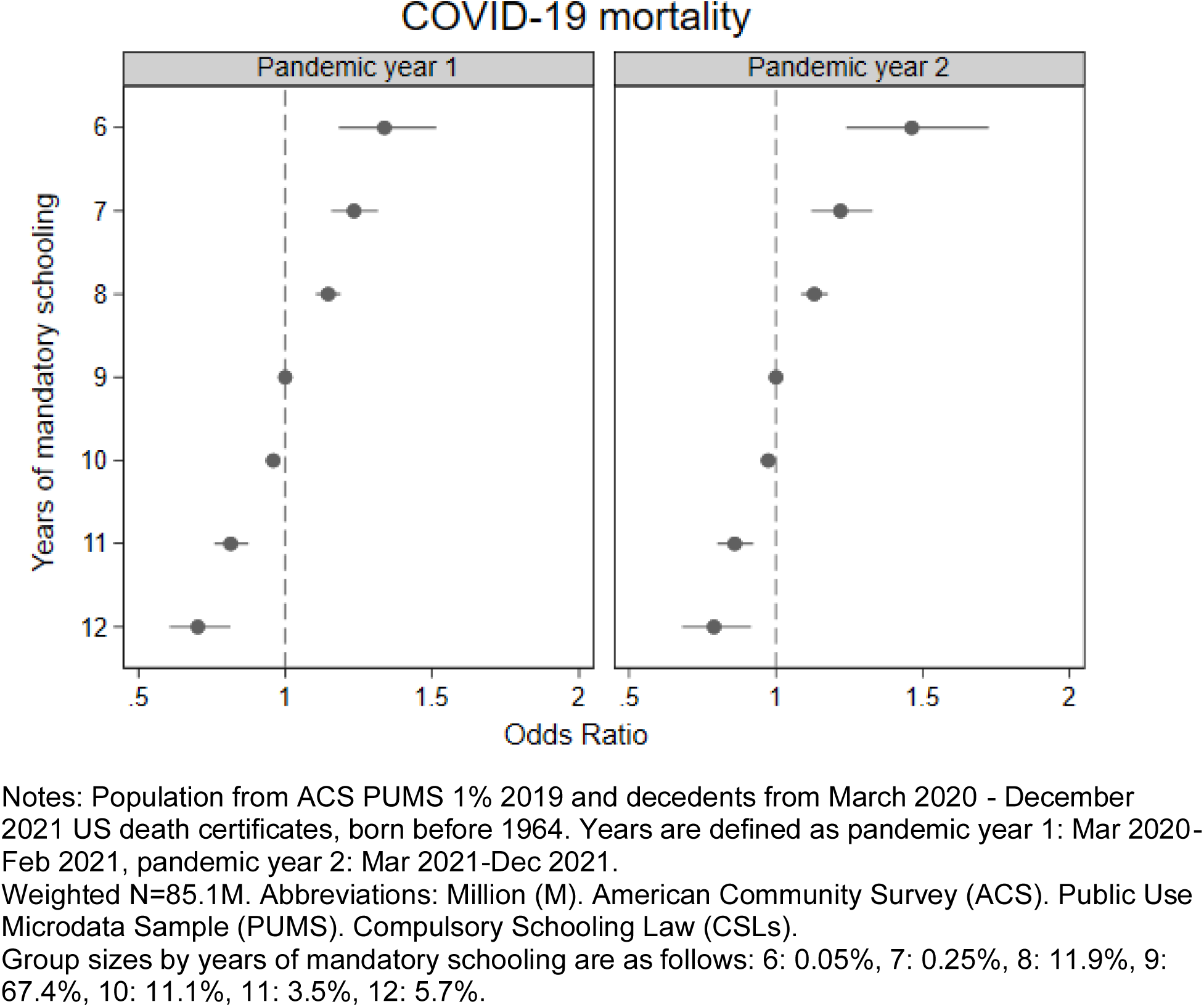
Effect of CSLs on COVID-19 mortality, Mar 2020-Dec 2021.

### Effect of CSLs on all-cause mortality

We observed a very similar dose-response relationship between CSLs and all-cause mortality, with no evidence of a differential effect when contrasting the pre-pandemic (year 0) and pandemic periods (years 1-2) (Figure 3). In the year prior to the pandemic, being mandated to complete 8 (vs 9) years of schooling was associated with higher odds of mortality (OR: 1.12; 95% CI: 1.09, 1.16), while 10 (vs 9) years was associated with lower odds (OR: 0.89; 95% CI: 0.87, 0.91).

**Figure 3.**
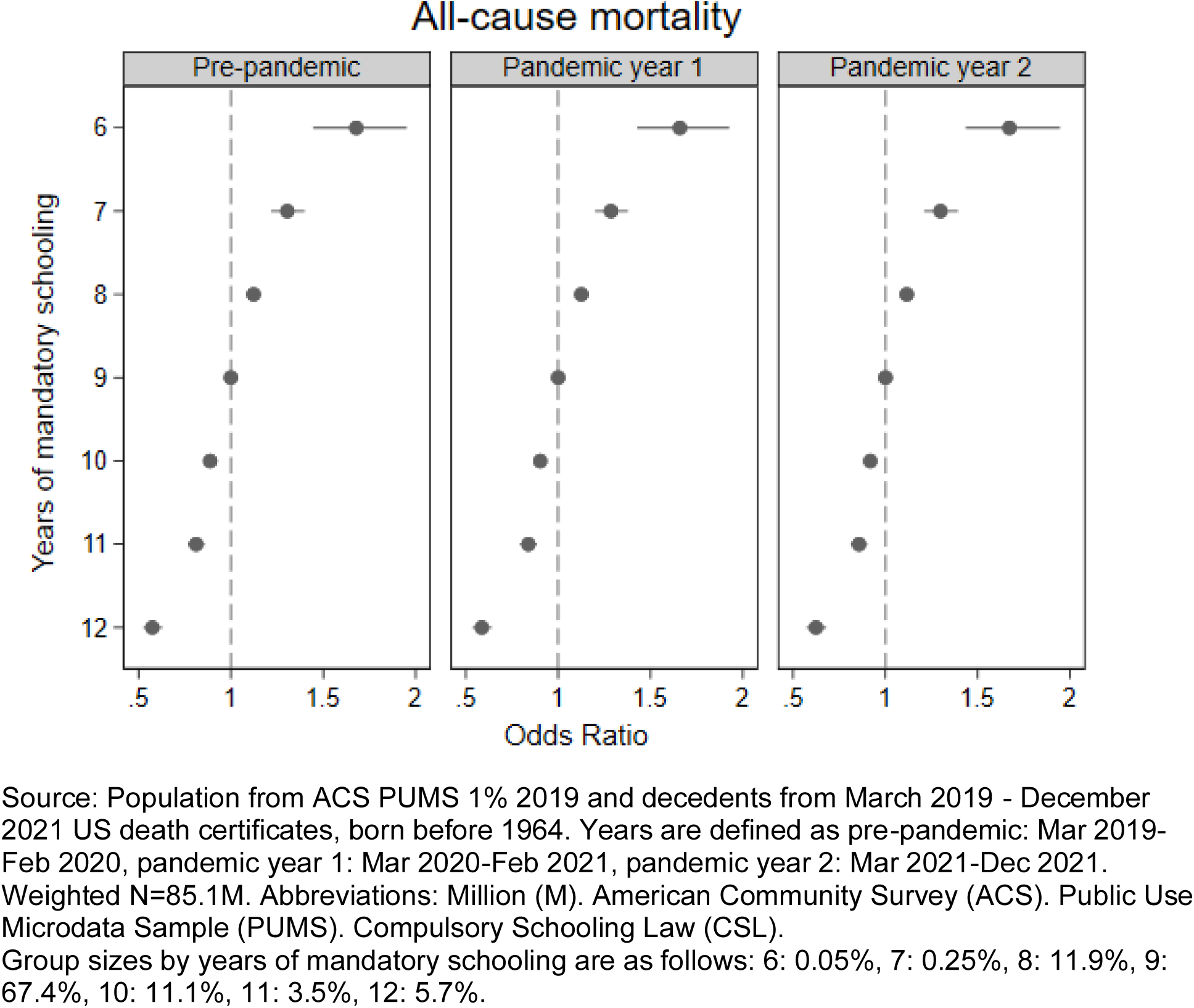
Effect of CSLs on all-cause mortality, Mar 2019-Dec 2021.

The corresponding estimates for all-cause mortality in the first and second year of the pandemic were similar. Being mandated to complete 8 (vs 9) years of schooling was associated with higher odds of mortality (OR: 1.13; 95% CI: 1.09, 1.16 first year; OR 1.12; 95% CI: 1.08 1.15, second year), while 10 (vs 9) years was again associated with lower odds (OR: 0.90; 95% CI: 0.88, 0.92 first year; OR: 0.92; 95% CI: 0.90, 0.94 second year). In all three time periods, individuals with less than 8 or more than 10 years of mandatory education represent relatively small groups, however, similar to the results for COVID-19 mortality, the dose-response gradient was present across all groups.

### Secondary analyses

We conducted a sensitivity analysis using an alternative definition of CSLs. Specifically, we assessed the effect of CSLs on COVID-19 and all-cause mortality using work permit age rather than legal drop-out age; while a similar overall trend appeared visible, the pattern in the years of data with the largest groups (7, 8, and 9 years of mandatory schooling) was relatively flat (details in Supplement).

We assessed potential effect modification of the relationship between CSLs and all-cause mortality by sex and race and ethnicity; overall trends did not differ substantially across subgroups; however further investigation is warranted (details in Supplement).

Since our mortality dataset was limited to December 2021, yielding study periods of unequal duration, we additionally examined an alternative specification of our time periods, using 10-month periods consistently for years 0, 1, and 2; results were very similar (details in Supplement).

## Discussion

Using national mortality data for the first two years of the pandemic and the year prior to the pandemic, we demonstrated that both COVID-19 and all-cause mortality were associated with mandated schooling requirements decades earlier. By incorporating state fixed effects, we demonstrate this is not merely a geographic pattern of COVID-19 mortality but that within each state, birth cohorts with longer required schooling experienced lower mortality. The clear dose-response gradient, with longer education mandates associated with lower mortality across the range of required schooling from 6 to 12 years, corroborated the relevance of educational context. The magnitude of associations was generally similar prior to the pandemic and during the pandemic.

Our findings are consistent with existing evidence that more years of attained education reduces mortality, including a 2024 meta-analysis finding a dose-response relationship between completed education and adult mortality globally.^22^ Our findings are also in line with prior evidence from Case and Deaton highlighting that the magnitude of the observational association between greater education and decreased all-cause mortality remained similar leading up to and during the COVID-19 pandemic.^23^ Our research builds on existing evidence by leveraging historical variation in CSLs and an ITT approach^24^ rather than traditional confounder-control approaches.

The results are also consistent with extensive evidence that education is a highly flexible resource individuals deploy to promote their health even in the context of rapid changes in the major threats to health. This is generally consistent with Fundamental Cause Theory.^11,12^ However, we did not find evidence to support our expectation that the educational inequality would grow as the pandemic evolved and a larger repertoire of preventive or treatment strategies became available.^12^ This may reflect the fact from essentially the first weeks of the pandemic, there was a known preventive strategy– physical isolation–differentially available to highly educated individuals. Against this backdrop of a highly effective preventive strategy for a very severe disease, additional evidence and tools for prevention may not have substantially increased the educational advantage.

Educational attainment is the most obvious mechanism linking mandated schooling and subsequent mortality. We hypothesize however that the school policies regarding enrollment and dropout age also changed within states in synchrony with other investments to improve educational attainment and quality, such as longer school term lengths, higher teacher pay, smaller class sizes, and greater investment in school resources.^8^ These investments may have enhanced the resources students attained from any given years of completed schooling, for example via improved literacy and numeracy. Although CSLs predict educational attainment, the associations are too small to plausibly fully account for the large effects of CSLs on mortality. Our analysis effectively rules out individual-level confounders such as childhood socioeconomic status or intellectual ability but cannot rule out other state policy changes that may have been adopted in tandem with increases in compulsory education.

We also cannot infer why education may influence mortality, although the observations suggest some mechanisms. The education benefit–in both observational studies and in the current study of education policies–extended to older adults unlikely to remain in the labor force. Thus, direct occupational exposures are likely not a full explanation of the educational inequality. Occupational exposures may indirectly affect mortality of older adults via co-resident working-age adults, however.^25^ Additional mechanisms include literacy/numeracy or the ability to sift through conflicting reports about the disease and its dangers. Institutional trust may also be relevant, especially as the pandemic became increasingly politically polarized. Financial security and lifelong accumulation of comorbid conditions prior to the pandemic may also have influenced educational patterns.

This research has several strengths. Our data sources provided a unique opportunity to link to CSLs given the availability of state and year of birth for every individual. Existing research has found somewhat mixed effects of CSLs using IV,^7^ potentially due to limited power or because of underlying heterogeneity in the impacts of increased schooling.^26^ Our research tried to address prior challenges by using a 1% population sample, all death certificate data, and ITT estimates. By combining ACS population data with the national vital statistics data, we simulated a national cohort. Additionally, the size of our dataset enabled relatively precise estimates.

This research also has several limitations. We restricted to individuals born before 1964; while CSLs may be less relevant for more recent birth cohorts, it would be valuable to understand how more recent educational policies designed to increase education (such as universal preschool or policies to increase college accessibility) have influenced health and mortality. Our research was also restricted to individuals born in the US, however policies aimed at increasing educational attainment may also have influenced individuals born outside the US; future work could assess the generalizability of our findings to wider populations. Finally, we cannot rule out potential state-level confounding factors such as co-occurring policy changes. However, our approach circumvents individual-level confounding factors, many of which are challenging to fully measure in existing research. Our results are therefore valuable for evidence triangulation regarding the relationship between education and mortality, by contributing insights that rest upon different assumptions than the majority of existing confounder-control research on this topic.

State policies regulating mandated schooling, implemented decades ago, continue to influence mortality of middle-aged and older adults who attended school under those policy regimes. Longer school mandates predict lower all-cause and COVID-19 mortality, likely via increased educational attainment and related educational experiences.

## Data Availability

The data used to estimate the population at-risk is available publicly via IPUMS. The restricted death certificate data is available via review and approval from the National Center for Health Statistics.

## References

1. Matthay EC, Duchowny KA, Riley AR, Thomas MD, Chen YH, Bibbins-Domingo K, et al. Occupation and Educational Attainment Characteristics Associated With COVID-19 Mortality by Race and Ethnicity in California. JAMA Netw Open. 2022;5(4):e228406–e228406.

2. Riley AR, Kiang MV, Chen YH, Bibbins-Domingo K, Glymour MM. Recent Shifts in Racial/Ethnic Disparities in COVID-19 Mortality in the Vaccination Period in California. J Gen Intern Med. 2022;37(7):1818–20.

3. Chen YH, Riley AR, Duchowny KA, Aschmann HE, Chen R, Kiang MV, et al. COVID-19 mortality and excess mortality among working-age residents in California, USA, by occupational sector: a longitudinal cohort analysis of mortality surveillance data. Lancet Public Health. 2022;7(9):e744–53.

4. Chen YH, Glymour MM, Catalano R, Fernandez A, Nguyen T, Kushel M, et al. Excess mortality in California during the coronavirus disease 2019 pandemic, March to August 2020. JAMA Intern Med. 2021;181(5):705–7.

5. Chen YH, Matthay EC, Chen R, DeVost MA, Duchowny KA, Riley AR, et al. Excess Mortality in California by Education During the COVID-19 Pandemic. Am J Prev Med. 2022;

6. Hawkins RB, Charles EJ, Mehaffey JH. Socio-economic status and COVID-19– related cases and fatalities. Public Health. 2020;189:129–34.

7. Hamad R, Elser H, Tran DC, Rehkopf DH, Goodman SN. How and why studies disagree about the effects of education on health: A systematic review and meta-analysis of studies of compulsory schooling laws. Soc Sci Med. 2018;212:168–78.

8. Brenowitz WD, Manly JJ, Murchland AR, Nguyen TT, Liu SY, Glymour MM, et al. State school policies as predictors of physical and mental health: A natural experiment in the REGARDS cohort. Am J Epidemiol. 2020;189(5):384–93.

9. Glymour MM, Kawachi I, Jencks CS, Berkman LF. Does childhood schooling affect old age memory or mental status? Using state schooling laws as natural experiments. J Epidemiol Community Health. 2008;62(6):532–7.

10. Hamad R, Nguyen TT, Bhattacharya J, Glymour MM, Rehkopf DH. Educational attainment and cardiovascular disease in the United States: a quasi-experimental instrumental variables analysis. PLoS Med. 2019;16(6):e1002834.

11. Clouston SA, Link BG. A retrospective on fundamental cause theory: State of the literature and goals for the future. Annu Rev Sociol. 2021;47:131–56.

12. Clouston SA, Natale G, Link BG. Socioeconomic inequalities in the spread of coronavirus-19 in the United States: A examination of the emergence of social inequalities. Soc Sci Med. 2021;268:113554.

13. Ruggles S, Flood S, Sobek M, Backman D, Chen A, Cooper G, et al. IPUMS USA: Version 15.0 [Internet]. Minneapolis, MN: IPUMS; 2024 [cited 2024 Feb 20]. Available from: https://usa.ipums.org

14. American Community Survey Office U.S. Census Bureau. American Community Survey 2021 1-year: PUMS User Guide and Overview. 2022;

15. Lleras-Muney A. Were Compulsory Attendance and Child Labor Laws Effective? An Analysis from 1915 to 1939. J Law Econ. 2002;45(2):401–35.

16. Acemoglu D, Angrist J. How Large are the Social Returns to Education? Evidence from Compulsory Schooling Laws. 1999;

17. Manly JJ, Murchland AR, Nguyen TT, Eng C, Doshi M, McClure LA, et al. Race and Place Specific Measures of School Quality: Validation and Relationship to Older Adult Health and Cognitive Function. Under Review.;

18. Commerce USDo. Statistical Abstract of the United States. Washington: US Department of Commerce, Bureau of the Census;

19. Irish AM. Education and later-life blood pressure: evidence from compulsory schooling laws and college expansion in the United States. 2023;

20. Card D, Krueger AB. Does school quality matter? Returns to education and the characteristics of public schools in the United States. J Polit Econ. 1992;100(1):1– 40.

21. COVID Data Tracker: Maps of COVID-19 Vaccinations by Age and Sex over Time: Percent of the Total Population with at Least One Dose of the COVID-19 Vaccine [Internet]. Centers for Disease Control and Prevention; Available from: https://covid.cdc.gov/covid-data-tracker/#vaccination-demographics-maps

22. Balaj M, Henson CA, Aronsson A, Aravkin A, Beck K, Degail C, et al. Effects of education on adult mortality: a global systematic review and meta-analysis. Lancet Public Health. 2024;

23. Case A, Deaton A. Mortality rates by college degree before and during COVID-19. National Bureau of Economic Research; 2021.

24. Chernozhukov V, Hansen C. The reduced form: A simple approach to inference with weak instruments. Econ Lett. 2008;100(1):68–71.

25. Brandén M, Aradhya S, Kolk M, Härkönen J, Drefahl S, Malmberg B, et al. Residential context and COVID-19 mortality among adults aged 70 years and older in Stockholm: a population-based, observational study using individual-level data. Lancet Healthy Longev. 2020;1(2):e80–8.

26. Lleras-Muney A. Education and income gradients in longevity: The role of policy. Can J Econ Can Déconomique. 2022;55(1):5–37.

