## Supplement for "Historical state compulsory schooling laws and pandemic-era mortality: A quasi-experimental study"

### Supplemental Analysis

Existing literature leveraging CSLs has often examined CSLs based both on legal drop-out age as well as work permit age. We therefore conducted an additional analysis including the CSLs based on work permit age (Supplemental Figure 2 & 3). Overall, the data appeared to show a dose-response gradient, however, in the years of mandatory schooling that contained the majority of the sample (7, 8, and 9 years), the results were flat. Overall, this suggests that the CSLs based on work permit age did not impact future mortality as strongly as CSLs based on legal drop-out age.

We additionally assessed both sex and racial identity as potential modifiers of the effects of CSLs on All-Cause mortality. Prior research has found that CSLs differentially affected both education and health outcomes for White and Black individuals, likely linked to school segregation during this period and differences in labor market opportunities.[1–3] Scholars have also suggested some differences in the causal effects of educational attainment on mortality by sex.[4] Based on this research, we assessed potential effect modification comparing White and Black individuals and comparing men and women in our sample. We examined stratified estimates for all-cause mortality across the full period. While interaction terms showed statistical evidence for interaction, the overall direction of the trend was similar when stratified by sex (Supplemental Figure 4) or race (Supplemental Figure 5). One result (12 years of mandatory schooling among Black individuals) deviates from the trend, but the confidence interval is extremely wide and overlaps the null. This analysis was exploratory and further investigation is warranted.

Given that 2022 death certificate data were not released at time of writing, our pandemic year 2 (Mar 2021 - Dec 2021) time period was two months shorter than our pre-pandemic (Mar 2019 - Feb 2020) and pandemic year 1 (Mar 2020 - Feb 2021) time periods. We therefore conducted a sensitivity analysis excluding the final two months from the pre-pandemic and pandemic year 1 periods to match the duration and months of the year in all our time periods. The results were very similar to our main analysis (Supplemental Figure 6 & 7).

### References


1. Brenowitz WD, Manly JJ, Murchland AR, Nguyen TT, Liu SY, Glymour MM, et al. State school policies as predictors of physical and mental health: A natural experiment in the REGARDS cohort. American journal of epidemiology. 2020;189(5):384–93.

2. Lleras-Muney A. Were Compulsory Attendance and Child Labor Laws Effective? An Analysis from 1915 to 1939. The Journal of Law & Economics. 2002;45(2):401–35.

3. Irish AM. Education and later-life blood pressure: evidence from compulsory schooling laws and college expansion in the United States. 2023;

4. Galama TJ, Lleras-Muney A, Van Kippersluis H. The effect of education on health and mortality: a review of experimental and quasi-experimental evidence. 2018;

**Supplemental Figure 1.** Conceptual diagram for the hypothesized effects of CSLs on mortality

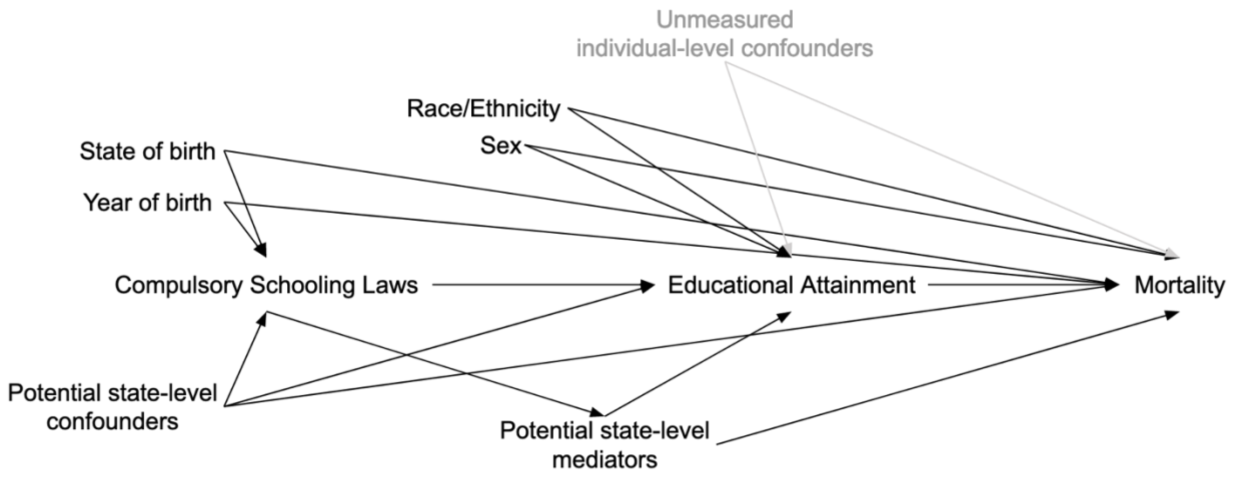


Notes: Unmeasured individual-level confounding pathways are shown in light gray, given these do not confound the effect of CSLs. Race/ethnicity and sex also do not confound the effect of CSLs, however we have included them in black as we have assessed potential effect modification in the effect of CSLs on mortality on these two factors. We have attempted to control for potential state-level confounders by adjusting for state-level characteristics at school-age.

Abbreviations: Compulsory Schooling Law (CSLs).

**Supplemental Figure 2.** Effect of CSLs on COVID-19 mortality, CSLs based on work permit age, Mar 2019-Dec 2021

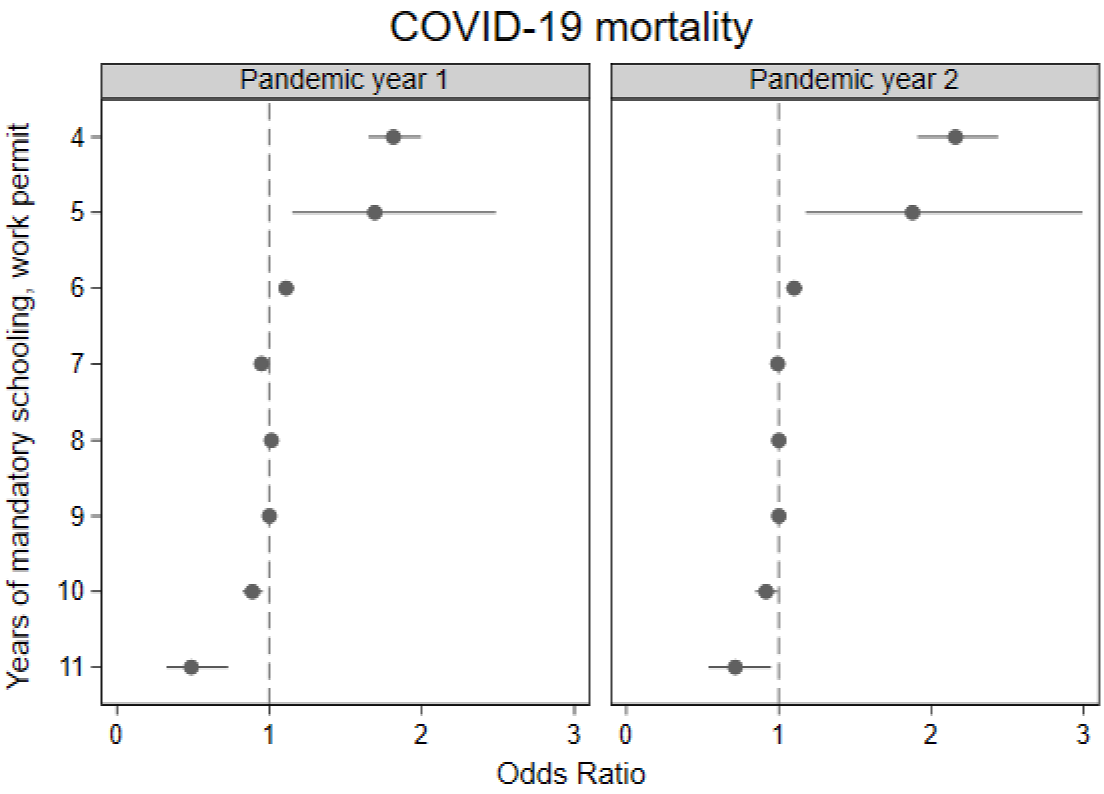


Notes: Population from ACS PUMS 1% 2019 and decedents from March 2019 - December 2021 US death certificates, born before 1964. Years are defined as pandemic year 1: Mar 2020-Feb 2021, pandemic year 2: Mar 2021-Feb 2022. Weighted N=85.1M. Abbreviations: Million (M). American Community Survey (ACS). Public Use Microdata Sample (PUMS). Compulsory Schooling Law (CSL).

**Supplemental Figure 3.** Effect of CSLs on all-cause mortality, CSLs based on work permit age, March 2019-December 2021

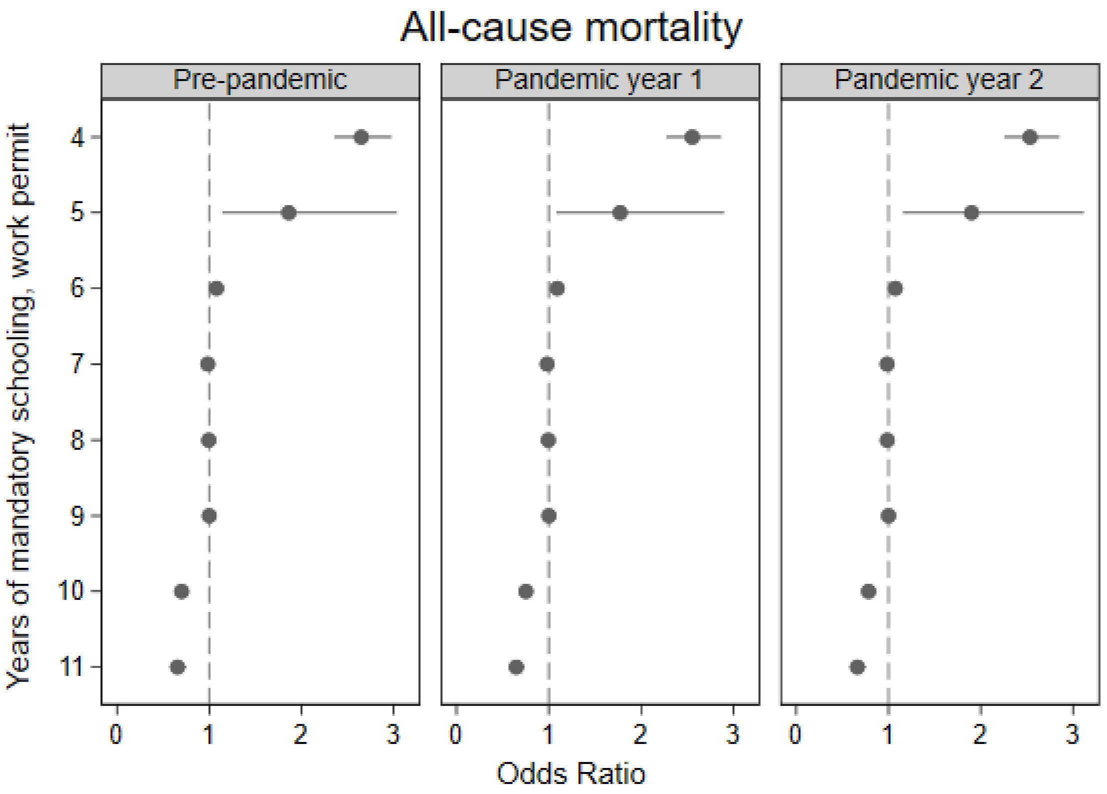


Notes: Population from ACS PUMS 1% 2019 and decedents from March 2019 - December 2021 US death certificates, born before 1964. Years are defined as pre-pandemic: Mar 2019-Feb 2020, pandemic year 1: Mar 2020-Feb 2021, pandemic year 2: Mar 2021-Dec 2021.

Weighted N=85.1M. Abbreviations: Million (M). American Community Survey (ACS). Public Use Microdata Sample (PUMS). Compulsory Schooling Law (CSL).

**Supplemental Figure 4.** Effect of CSLs on all-cause mortality, stratified by sex, Mar 2020-Dec 2021

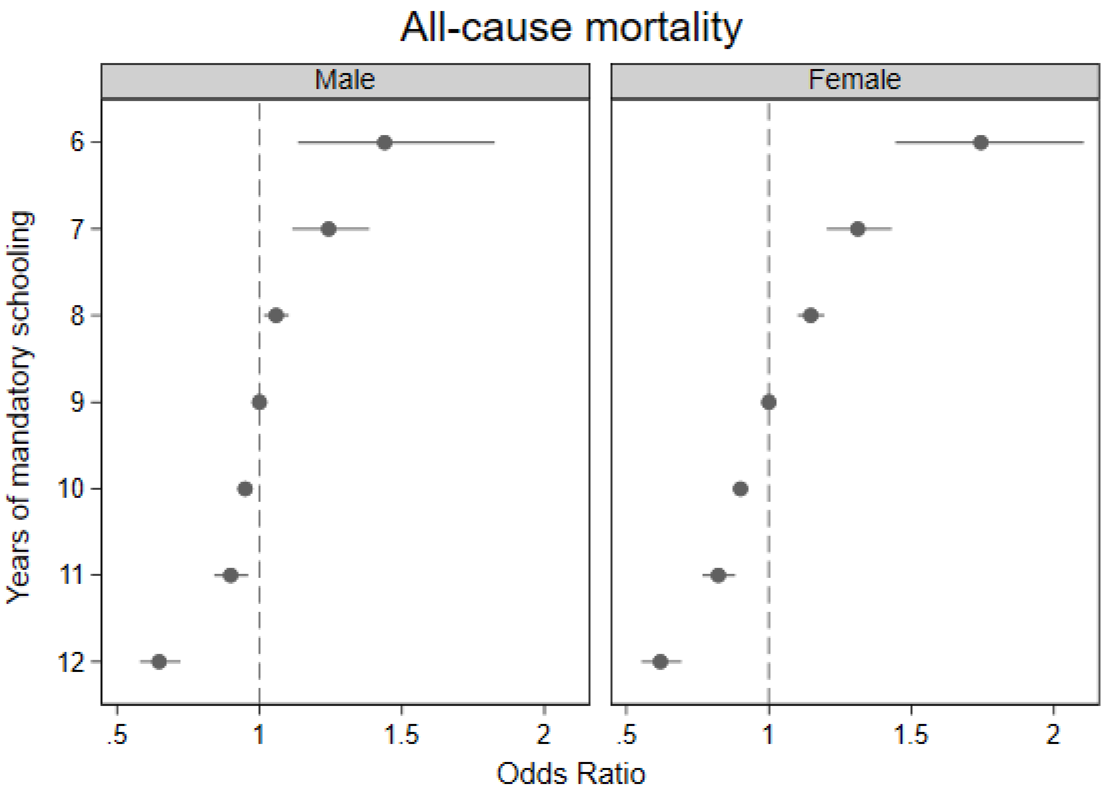


Notes: Population from ACS PUMS 1% 2019 and decedents from March 2020 - December 2021 US death certificates, born before 1964.
Weighted N=85.1M. Abbreviations: Million (M). American Community Survey (ACS). Public Use Microdata Sample (PUMS). Compulsory Schooling Law (CSL).

**Supplemental Figure 5.** Effect of CSLs on all-cause mortality, stratified by race/ethnicity, Mar 2020-Dec 2021

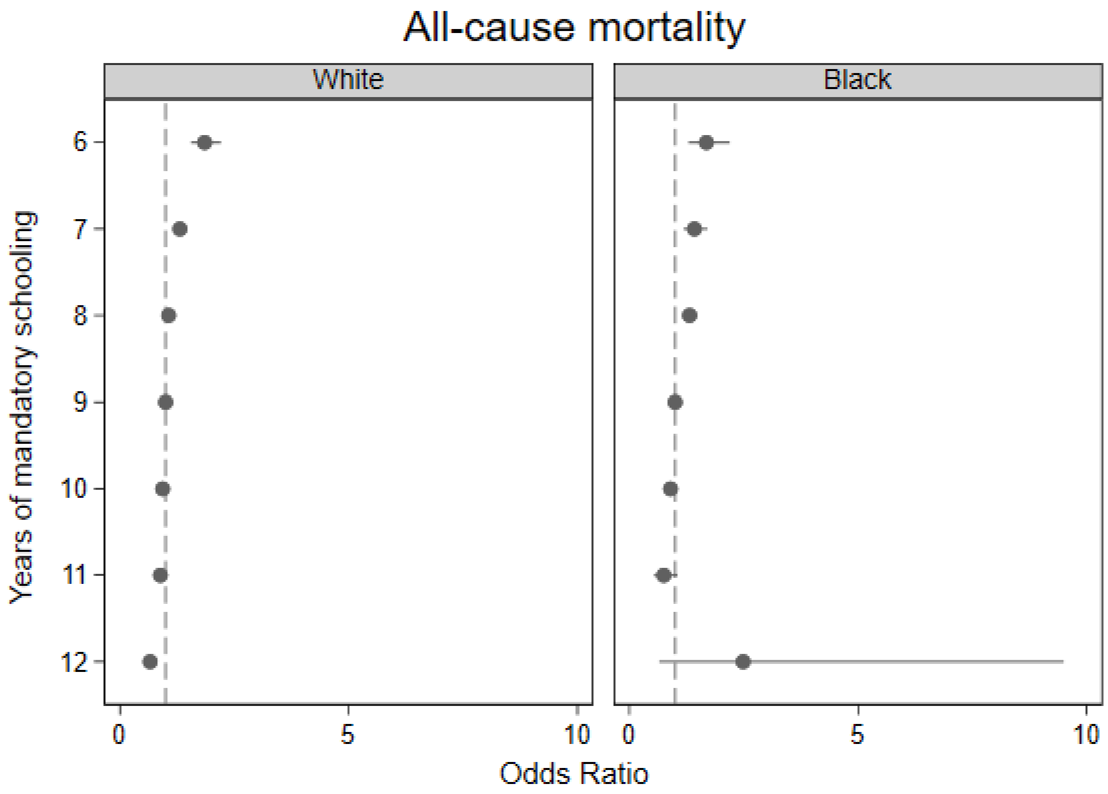


Notes: Population from ACS PUMS 1% 2019 and decedents from March 2020 - December 2021 US death certificates, born before 1964.
Weighted N=85.1M. Abbreviations: Million (M). American Community Survey (ACS). Public Use Microdata Sample (PUMS). Compulsory Schooling Law (CSL).

**Supplemental Figure 6.** Effect of CSLs on COVID-19 mortality, adjusted time periods, Mar 2020-Dec 2021

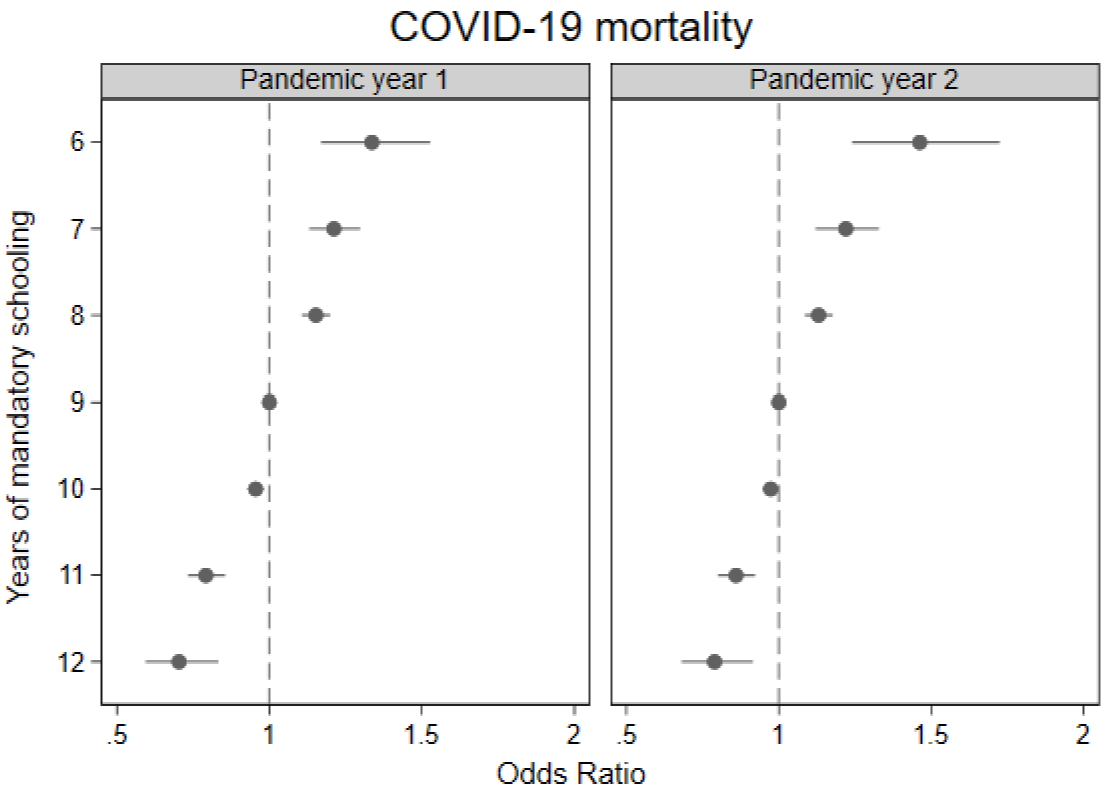


Notes: Population from ACS PUMS 1% 2019 and decedents from March 2020 - December 2021 US death certificates, born before 1964. Excludes final two months of pandemic year 1 period to match unavailable months in pandemic year 2 period. Years are defined as pandemic year 1: Mar 2020-Dec 2020, pandemic year 2: Mar 2021-Dec 2021.

Weighted N=84.2M. Abbreviations: Million (M). American Community Survey (ACS). Public Use Microdata Sample (PUMS). Compulsory Schooling Law (CSL).

**Supplemental Figure 7.** Effect of CSLs on all-cause mortality, adjusted time periods, Mar 2019-Dec 2021

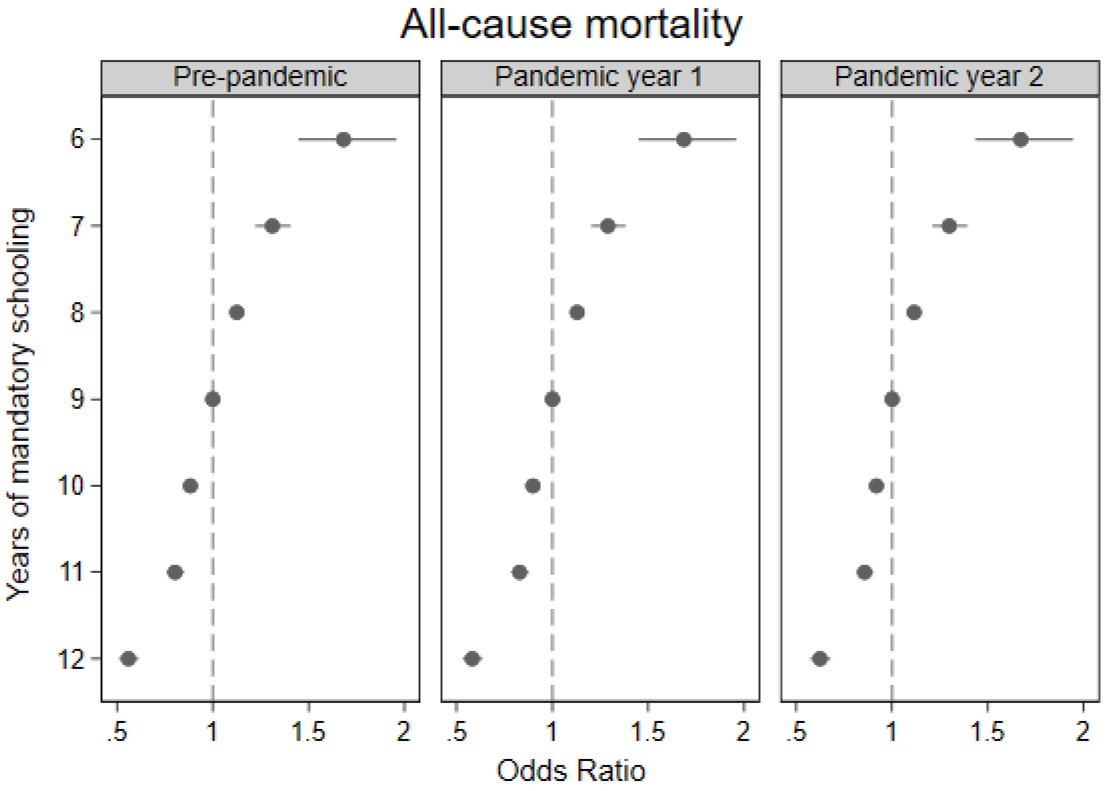


Notes: Population from ACS PUMS 1% 2019 and decedents from March 2019 - December 2021 US death certificates, born before 1964. Excludes final two months of pre-pandemic and pandemic year 1 periods to match unavailable months in pandemic year 2 period. Years are defined as pre-pandemic: Mar 2019-Dec 2019, pandemic year 1: Mar 2020-Dec 2020, pandemic year 2: Mar 2021-Dec 2021.
Weighted N=84.2M. Abbreviations: Million (M). American Community Survey (ACS). Public Use Microdata Sample (PUMS). Compulsory Schooling Law (CSL).

**Supplemental Table 1.** Risk of mortality in the target population during each time period

| | Mar 2019-Feb 2020 | Mar 2020-Feb 2021 | Mar-Dec 2021 |
|---|---|---|---|
| COVID-19 mortality | - | 0.41% | 0.23% |
| All-Cause mortality | 2.48% | 2.99% | 2.41% |

Notes: Population from ACS PUMS 1% 2019 and decedents from March 2019 - December 2021 US death certificates, born before 1964.
Weighted N=85.1M. Abbreviations: Million (M). American Community Survey (ACS). Public Use Microdata Sample (PUMS).
